# A Reproducible Framework for Integrating Chronic Deep Brain Stimulation Sensing with Wearable Behavioral Monitoring

**DOI:** 10.64898/2026.09.18.26363453

**Authors:** Saipravallika Chamarthi, Gabriel Reyes, Tomasz M. Fraczek, Sophia Pouya, Yewen Zhou, Thomas P. Kutcher, Rick R. Hanish, Ashley Deng, Jeffrey A. Herron, Eric A. Storch, Wayne K. Goodman, Sameer A. Sheth, Nicole R. Provenza

## Abstract

**Objective:** Chronic sensing-enabled deep brain stimulation (DBS) devices enable long-term neural recordings in naturalistic settings, but interpreting these data requires concurrent behavioral context. Our objective was to develop a reproducible end-to-end framework for continuous acquisition and synchronization of wearable-derived behavioral data alongside chronic DBS recordings to enable longitudinal neurobehavioral studies.

**Approach:** We developed a publicly available software framework that includes automated wearable data ingestion, neural artifact handling, epoch-based temporal synchronization, and generation of analysis-ready neurobehavioral datasets. We tested this framework using simultaneous sensing-enabled DBS and Oura Ring recordings from three participants with obsessive-compulsive disorder.

**Main results:** Using this framework, we synchronized 6,384 hours of intracranial neural recordings and wearable-derived data. The resulting multimodal neurobehavioral datasets spanned months of ambulatory monitoring and integrated chronic neural recordings with sleep-wake state, physical activity, autonomic physiology, and DBS parameters. Benchtop testing revealed 12 seconds of Medtronic Percept clock drift relative to network time over a one-week period. This temporal error was substantially smaller than the 10-minute sampling interval of the chronic neural recordings, supporting reliable alignment with wearable-derived data.

**Significance:** This work provides an open-source, reproducible framework for continuous acquisition, synchronization, and analysis of wearable-derived data alongside sensing-enabled DBS recordings. By reducing the technical barriers to generating behaviorally annotated neural datasets, this framework enables scalable longitudinal neurobehavioral studies and provides practical foundation for biomarker discovery.

## INTRODUCTION

Understanding how neural activity relates to behavior is central to the development of clinically useful neurophysiological biomarkers. Traditionally, this relationship has been studied during structured laboratory or clinical assessments, where neural recordings are synchronized with behavioral tasks or external measurements such as video and audio.^1,2^ Although these controlled settings provide highly interpretable data, they capture only brief snapshots of behavior and may fail to reflect the slow, long-timescale, state-dependent dynamics that characterize many chronic neurological and psychiatric disorders.^3^ Ambulatory monitoring therefore offers an opportunity to study neurobehavioral dynamics continuously in the environments where symptoms naturally occur.

Deep brain stimulation (DBS) has transformed the feasibility of ambulatory intracranial monitoring through sensing-enabled implantable pulse generators (IPGs) capable of continuously recording local field potential (LFP) activity during daily life.^4^ Although DBS programming continues to rely on intermittent clinical visits, chronic sensing enables longitudinal and continuous tracking of neural features over clinically relevant timescales. Without concurrent behavioral context, however, these recordings remain difficult to interpret. This challenge is particularly salient in psychiatric DBS, where therapeutic effects are often delayed and symptom trajectories are heterogeneous. In treatment-resistant obsessive-compulsive disorder (OCD), for example, meaningful improvement may lag behind stimulation parameter changes by weeks to months, leaving clinicians dependent on intermittent rating scales and patient self-report to guide therapy.^5^ These limitations motivate approaches that continuously capture objective behavioral context alongside chronic neural recordings.

Consumer wearables have expanded the ability to measure behavior continuously in naturalistic settings through passive monitoring of sleep, physical activity, autonomic physiology, and other clinically relevant processes.^6^ Modern wearable devices provide reliable estimates of these behavioral states, enabling scalable digital phenotyping outside the clinic.^7^ Importantly, these measurements offer the behavioral context needed to interpret chronic neural recordings, allowing fluctuations in brain activity to be related to objectively measured changes in daily life. Together, these advances motivate a convergent strategy for psychiatric neuromodulation in which wearable-derived state markers are coupled with chronic intracranial features to interpret symptom dynamics in context and to inform personalized or adaptive stimulation strategies.

Despite these complementary advances, integrating chronic neural recordings with wearable-derived behavioral data remains challenging because the two systems operate independently, differ in sampling strategies, and maintain separate clocks. Here, we present a reproducible end-to-end framework for continuous wearable data acquisition, temporal synchronization with chronic DBS recordings, and generation of analysis-ready neurobehavioral datasets. Although developed and evaluated using the sensing-enabled Medtronic Percept DBS device and Oura Ring recordings in participants with OCD, the framework is broadly applicable to studies integrating chronic neural recordings with objective behavioral and physiological measurements and provides scalable infrastructure for longitudinal neurobehavioral research and multimodal biomarker discovery.

## METHODS

### Participants and Study Overview

Three participants with treatment-resistant OCD underwent DBS implantation with the Medtronic Percept PC system (NCT05915741). Medtronic SenSight leads were implanted bilaterally in the ventral capsule/ventral striatum (VC/VS) as described previously^8^, with the deepest contact positioned near the VC/VS white matter boundary. Each participant received an Oura Ring and subscription to the Oura application and was instructed to wear the device for at least 15 hours per day. All participants provided written informed consent under a protocol approved by the Institutional Review Board at Baylor College of Medicine (H-43144).

### Data Acquisition and Preprocessing

We collected chronic neural recordings using the Percept BrainSense Timeline modality, which provides 10-minute averages of local field potential (LFP) power in a user-defined 5 Hz-wide frequency band. We configured the BrainSense Timeline capability of the DBS devices to record VC/VS LFP power in a theta-alpha band centered near 9 Hz (8.79 ± 2.5 Hz, hereafter referred to as “9 Hz power”), along with the stimulation amplitude settings applied during active therapy. Participants could adjust stimulation amplitude within a clinician-defined range using a patient controller and were advised to reduce stimulation amplitude at night to facilitate sleep onset. Because the Percept PC retains chronic sensing data onboard for a limited duration (∼60 days), we offloaded data at scheduled clinical visits and transferred them to a secure institutional server.

To enable scalable longitudinal wearable data acquisition, we developed a near-automated ingestion workflow using the Oura Ring application programming interface (API; Figure 1). We registered each participant’s Oura Ring using a study-generated email address and authorized it via a one-time OAuth2 flow.^9^ We securely stored the resulting access token and refreshed it automatically every 12 hours to ensure uninterrupted data access.

**Figure 1.**
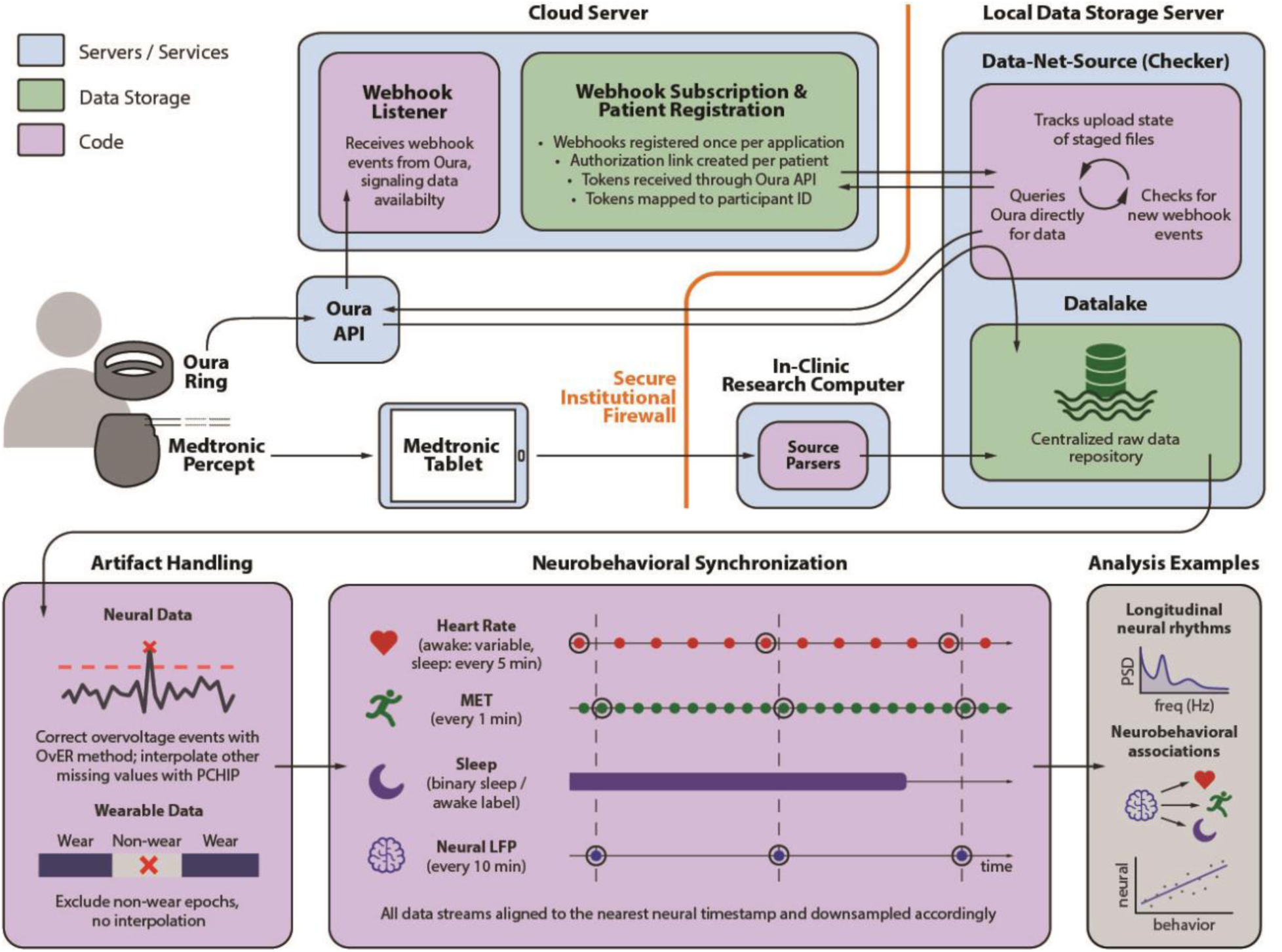
Data Synchronization Framework for Oura Ring and Medtronic Percept Metrics. Overview of the end-to-end neurobehavioral synchronization workflow, including data ingestion, artifact handling, temporal alignment, and example analyses enabled by the resulting datasets. Oura Ring data are detected as they become available through webhook subscriptions hosted on AWS, then retrieved through the Oura API by a local checker service and stored securely on our local data storage server. Neural data are manually offloaded from each participant’s implantable pulse generator (IPG) during clinical visits and uploaded to the study data server. During preprocessing, neural outliers are removed using Overvoltage Event Removal (OvER) and reconstructed with piecewise cubic Hermite interpolating polynomial (PCHIP) interpolation, while wearable non-wear periods are excluded and represented as NaN. All data streams are then temporally synchronized to the nearest neural data timestamp to facilitate longitudinal neurobehavioral analyses. MET: metabolic equivalent; PSD: power spectral density.

Wearable data ingestion proceeded in two sequential steps. First, we deployed an independent HTTPS listener on an Amazon Web Services Elastic Compute Cloud (AWS EC2) instance. We implemented the listener as a Flask web application and deployed it using Gunicorn. The listener subscribed to webhook event notifications across 16 physiological and behavioral data streams, including sleep architecture, physical activity, heart rate, oxygen saturation, stress, resilience, and workout metrics. We stored incoming payloads as structured JSON files organized by participant identifier and data type. Second, a downloader service running on our primary data server periodically queried the EC2 instance for new events and retrieved those newly available records, updating the local study data store for downstream preprocessing and synchronization.

To mitigate neural artifacts, we identified and corrected any overvoltage events using Overvoltage Event Removal (OvER), as described previously.^10^ Residual missing values were reconstructed using shape-preserving piecewise cubic Hermite interpolating polynomial (PCHIP) interpolation, with a maximum interpolated gap of 12 samples (2 hours).

### Temporal Standardization and Neurobehavioral Synchronization

We parsed all neural and wearable timestamps as time zone-aware datetimes and converted them to Unix epoch time (seconds since 1970-01-01 UTC) before synchronization. This common epoch-based time base standardized timestamp formats across the devices and avoided ambiguity introduced by time zone changes or daylight-saving time transitions. We converted epoch timestamps to local time for visualization and definition of local-day boundaries, but all synchronization operations remained in epoch time.

Because Oura Ring data are reported as interval-based measurements with only start and end datetimes, we expanded each data stream into explicitly timestamped observations according to its native temporal resolution (e.g., 5-minute sleep hypnograms and nocturnal physiology; 1-minute metabolic equivalent [MET] values). We used recorded sleep episodes and daily activity records to define the temporal windows available for alignment with neural data for each wearable-derived data stream. We then used chronic DBS recordings as the temporal reference for synchronization and assigned the nearest wearable observation to each neural timestamp only when that timestamp fell within the corresponding valid interval (e.g., sleep-stage labels during recorded sleep episodes, MET values during daily activity periods). This constraint prevented spurious nearest-neighbor matching across extended gaps in wearable data. The resulting synchronized dataset contained 10-minute LFP power estimates together with temporally matched sleep staging, MET, heart rate, heart rate variability (HRV), and other available behavioral and physiological measures.

We treated MET values of 0.1 as indicators of Oura Ring non-wear and set these values to missing to distinguish data collection gaps from physiological variation.^11,12^ We classified each synchronized timestamp as “worn” if either a sleep-stage label or valid MET value was available and “not worn” otherwise. We retained missing wearable values without imputation to preserve gaps due to non-wear or incomplete synchronization. Total overlapping hours reflected the summed duration of synchronized 10-minute epochs with concurrent neural and wearable-derived data.

### Benchtop Validation of Neural Timestamps

We performed a benchtop assessment of the Percept IPG clock drift to verify the accuracy of recorded neural timestamps. We synchronized the IPG to network time using the Medtronic Percept Clinician Programming Tablet by first connecting the tablet to Wi-Fi and then selecting “Update Device Time.” We disconnected the IPG from the tablet for approximately one week and withheld recharging. At the end of this period, we downloaded neural data immediately before and after re-synchronizing the device clock to network time using the “Update Device Time” function available on the tablet. We calculated the cumulative clock drift as the difference between the original and corrected timestamps assigned to each LFP value.

### Derived Measures and Statistical Analyses

We computed wearable-derived sleep duration and bedtime onset from Oura-determined sleep episode records and classified each 10-minute LFP power value as sleep or wake according to the synchronized wearable sleep-stage label. We then identified the daily 9 Hz power peak as the maximum value within each local midnight-to-midnight period. Because time of day is circular, we visualized peak timing using polar plots with the 24-hour cycle mapped onto 0–2π radians, preserving temporal adjacency across midnight. We estimated 95% confidence intervals around the observed circular median peak time from the distribution of circular medians obtained across 1,000 bootstrap resamples with replacement.

### Code Availability

We have made all software developed for wearable data acquisition, synchronization, and downstream neurobehavioral analysis publicly available. The Oura webhook listener and data downloader operate together to support near-automated longitudinal wearable data acquisition: the listener receives notifications when new Oura data become available, and the downloader retrieves these records and updates the local study data store. These repositories are available at https://github.com/BCM-Neurosurgery/oura-webhook-listener and https://github.com/BCM-Neurosurgery/oura-data-downloader, respectively. We implemented downstream preprocessing and synchronization in Python using a separate analysis framework that ingests Oura and Percept data, standardizes timestamps to Unix epoch time, performs temporal alignment, and generates analysis-ready neurobehavioral datasets. This repository is available at https://github.com/BCM-Neurosurgery/OuraRing-Pipeline-and-Utility.

All three software repositories are distributed under the same Baylor College of Medicine license, which permits noncommercial use by researchers at academic and nonprofit research institutions and requires acknowledgement through citation of this work. Commercial use requires separate permission from Baylor College of Medicine.

## RESULTS

### Longitudinal synchronization of ambulatory neural and wearable data

Across three participants implanted with sensing-capable DBS for treatment-resistant OCD, we synchronized 6,384 hours of overlapping chronic neural recordings and wearable-derived behavioral and physiological data using the end-to-end framework shown in Figure 1. The framework integrated near-automated Oura Ring acquisition, Percept data ingestion, neural artifact handling, temporal standardization, and multimodal alignment to generate analysis-ready neurobehavioral datasets. Daily wearable adherence exceeded the target of 15 hours per day across all participants (Fig. 2A), enabling dense longitudinal characterization of sleep, physical activity, autonomic physiology, and DBS therapy over months of ambulatory monitoring (Fig. 2A-F).

**Figure 2.**
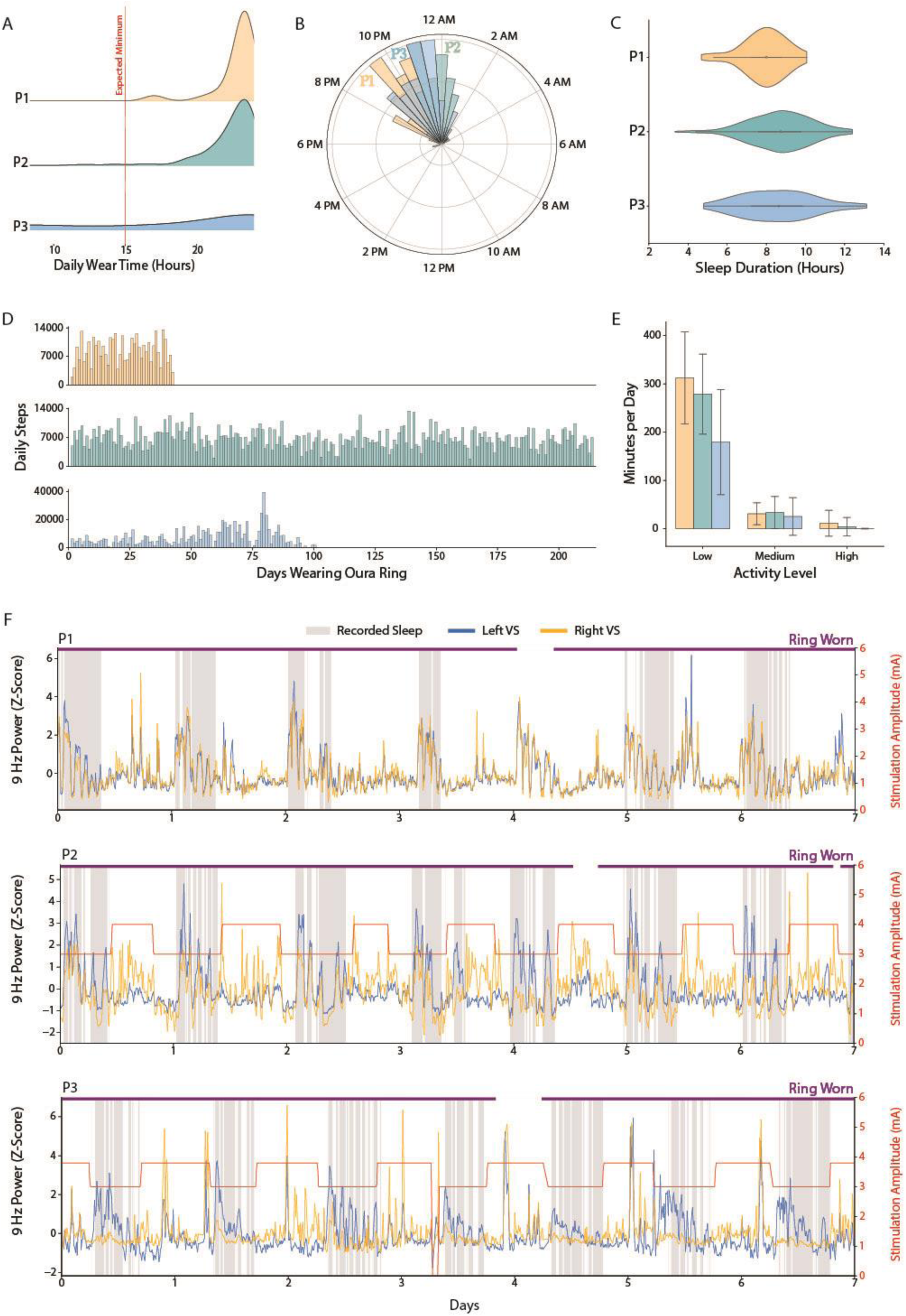
Synchronization of Oura Ring data with longitudinal neural recordings from Percept DBS system. (A–E) Representative longitudinal wearable measurements demonstrate high participant adherence and continuous monitoring of sleep, activity, and other daily behavior throughout ambulatory recording. (F) Representative synchronized recordings illustrate temporal alignment of bilateral chronic VC/VS LFP power, wearable-derived sleep intervals, stimulation amplitude, and device wear status on a common timeline, demonstrating the feasibility of generating multimodal neurobehavioral datasets suitable for longitudinal analysis. Stimulation amplitude is not shown for P1 because these recordings were collected prior to DBS activation.

Representative synchronized recordings in Fig. 2F illustrate the multimodal neurobehavioral dataset, aligning chronic neural activity, wearable-derived behavioral measures, and DBS parameters on a shared temporal axis. These recordings preserve the temporal relationships among sleep intervals, stimulation amplitude, wearable coverage, and chronic LFP activity over extended ambulatory monitoring periods. In participants who adjusted stimulation amplitude throughout the day, the synchronized recordings additionally capture the temporal relationship between behavioral state, patient-driven therapy adjustments, and chronic neural activity. Monitoring periods can additionally be aligned to DBS onset (t=0) to directly compare pre- and post-activation data across participants.

To validate timestamp accuracy, we performed a benchtop assessment of Medtronic Percept IPG clock drift. After approximately one week without network synchronization, the device exhibited a cumulative clock offset of 12 seconds relative to network time, corresponding to a constant shift across all recorded timestamps. This drift was substantially smaller than the 10-minute temporal resolution of the chronic sensing data, confirming that native Percept timestamps provide sufficient accuracy for reliable longitudinal synchronization.

### Preliminary neurobehavioral analyses enabled by synchronized datasets

To demonstrate the analytical capabilities of the synchronized dataset, we performed preliminary neurobehavioral characterizations that depended on reliable temporal alignment between Oura Ring state labels and chronic DBS feature streams. Temporal alignment between Oura-derived sleep staging and chronic DBS recordings enabled objective behavioral annotation of every neural sample, allowing state-dependent distributions of LFP activity to be examined over months of ambulatory monitoring (Fig. 3A). Large numbers of synchronized observations were available across participants and hemispheres despite intermittent changes in sensing availability associated with DBS programming configurations. These state-tagged distributions illustrate how wearable sleep labeling can provide scalable, objective behavioral context for chronic neural features without reliance on subjective bedtime reports.

**Figure 3.**
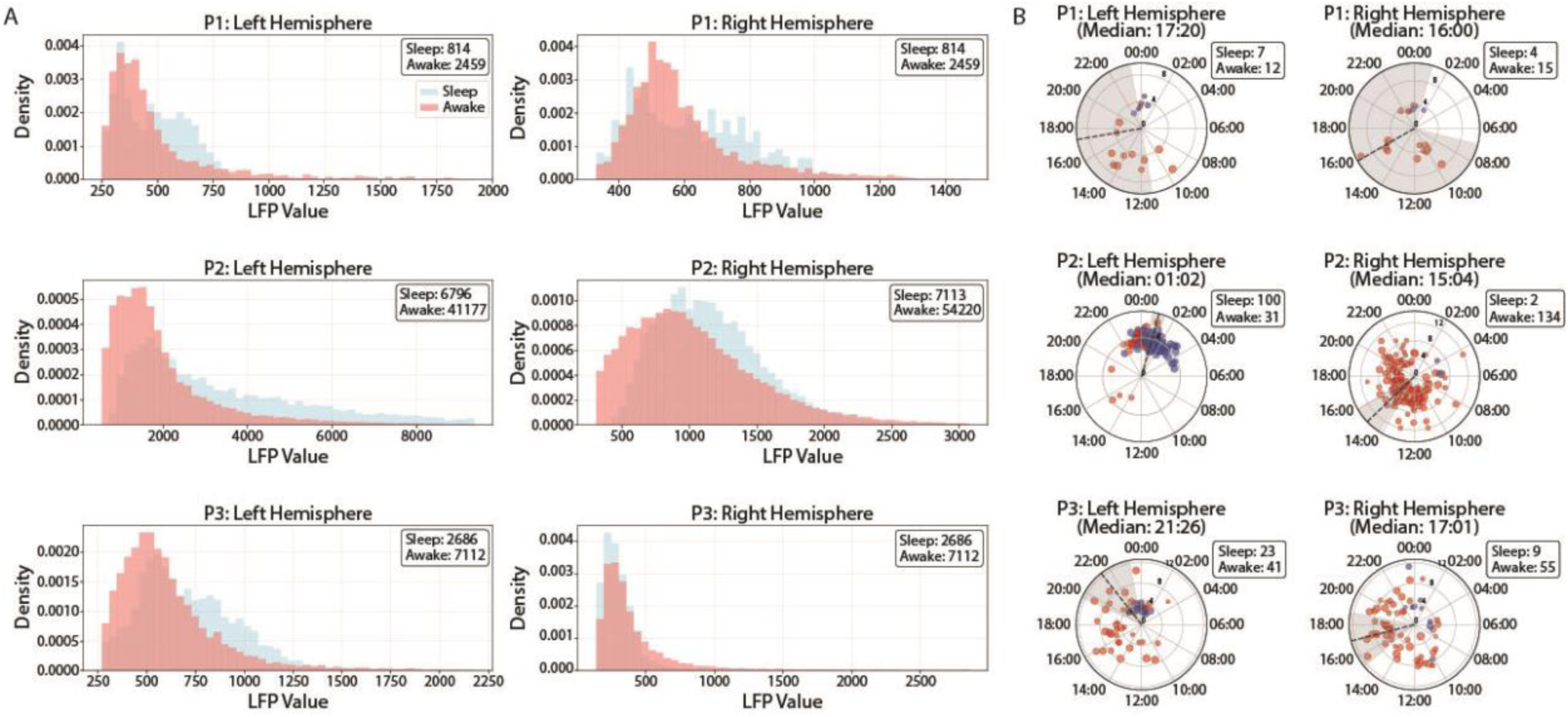
Longitudinal neurobehavioral analyses enabled by synchronized wearable and chronic DBS recordings. (A) Temporal alignment of wearable-derived sleep staging with chronic DBS recordings enables state-dependent comparisons of VC/VS 9 Hz power during sleep and wakefulness across participants. (B) Daily peak analyses identify the timing and behavioral state associated with maximum daily 9 Hz power, illustrating how synchronized behavioral annotations (sleep = blue, awake = red) facilitate characterization of longitudinal neural dynamics. The dotted black line and shaded gray region indicate the median time and 95% confidence interval, respectively. The number of recorded daily LFP peaks during sleep and wakefulness is denoted next to each polar plot. LFP peaks predominantly occurred during wakefulness in both hemispheres of P1 and P3; however, they were split between sleep (left hemisphere) and awake (right hemisphere) periods in P2.

To further demonstrate the utility of these synchronized behavioral annotations, we performed an exemplar analysis to characterize the temporal organization of chronic 9 Hz activity. We identified the timing of each day’s maximum neural power and classified each peak as occurring during sleep or wake using the synchronized behavioral labels. Peak timing varied across participants and hemispheres, with daily maxima occurring during either sleep or wake depending on the individual and recording hemisphere (Fig. 3B). In P2, the left hemisphere peaks occurred more frequently during sleep than wake, whereas right hemisphere peaks occurred predominantly during wake. In P1 and P3, however, peaks were predominantly classified during wake in both hemispheres. These results illustrate how synchronized neurobehavioral data may help disentangle the behavioral and temporal dynamics underlying neural features observed over long-term ambulatory recordings.

Finally, we evaluated relationships between wearable-derived physical activity (MET) and chronic 9 Hz power to demonstrate integration of neural and behavioral measures. Wearable-derived activity showed a weak positive association with 9 Hz LFP power across all three participants.

Collectively, these examples illustrate how synchronized neural and behavioral measurements enable quantitative characterization of longitudinal neurobehavioral dynamics beyond what is possible using either modality alone.

## DISCUSSION

Combining chronic neural recordings with wearable-derived behavioral measures enables neural activity to be interpreted in the context of everyday behavior. However, integrating these data streams requires synchronization across independent devices that differ in sampling frequencies and maintain separate clocks. Ambulatory sensing studies commonly rely on manual export of wearable data or custom laboratory-specific scripts.^3,10,13–15^ Our framework provides an openly available solution for near-automated acquisition and synchronization of longitudinal wearable data. The workflow leverages near-automated wearable data retrieval and native device timestamps to generate time-aligned multimodal datasets with minimal investigator effort. This framework can be readily adapted across studies, enabling scalable and reproducible analyses of naturalistic brain-behavior relationships.

The reliability of this synchronization strategy depended on accurate timekeeping onboard both devices. We therefore assessed the stability of the Percept PC internal clock in benchtop testing, observing 12 seconds of cumulative drift over one week. Since our protocol involves routine download of Percept recordings and synchronization of Percept clock with network time during clinical visits, if drift accumulated linearly, the anticipated offset would remain substantially smaller than the 10-minute sampling interval of the synchronized dataset. In other words, after 60 days of BrainSense Timeline recordings (the maximum duration before overwriting), total anticipated drift is less than 2 minutes. These findings demonstrate that native Percept timestamps are sufficiently stable for longitudinal multimodal synchronization, eliminating the need for external timing hardware or manual post hoc correction for applications at the temporal resolution of chronic BrainSense recordings.

A major application of this framework is continuous neurobehavioral phenotyping during chronic DBS therapy research studies. In psychiatric DBS, where therapeutic response often unfolds over weeks to months, objective behavioral monitoring may provide an important complement to intermittent clinician-administered assessments by capturing disease dynamics continuously throughout treatment. Even in movement disorders, these measures may provide insight into both motor and nonmotor symptom changes that influence quality of life as disease progresses. For instance, sleep disturbances are a hallmark of many neurological and neuropsychiatric disorders and are closely linked to circadian regulation, cognitive function, and symptom severity.^16–19^ Oura Ring-derived sleep staging can thus provide objective annotation of sleep architecture across months of ambulatory monitoring, allowing both transient and longitudinal changes in neural activity to be disentangled beyond the logistical constraints of polysomnography and patient sleep logs. Beyond sleep, behavioral and physiological processes, such as physical activity, heart rate, and heart rate variability, may reveal distinct dimensions of illness that would likely be obscured by aggregate symptom scores or evolve independently during treatment. Therefore, objective behavioral phenotyping can improve interpretation of chronic neural signals and facilitate discovery of clinically meaningful neural biomarkers.

Although intended primarily as demonstrations of the synchronized dataset, these preliminary analyses demonstrate the types of neurobehavioral questions that become feasible once chronic neural recordings are continuously annotated with objective behavioral state. For instance, apparent time-of-day structure in neural activity may reflect recurring behaviors that tend to occur at particular times, rather than time of day itself. Synchronized behavioral annotation can therefore help distinguish whether such neural motifs track behavioral state, are tied to a particular phase of day, or vary with changes in an individual’s daily routine. Rather than relying on isolated clinic visits or patient recall, investigators can use this framework to characterize state-dependent neural dynamics, examine circadian organization of neural activity, and relate chronic electrophysiological changes to objectively measured behavior over months of therapy.

Finally, although this framework was developed and validated using the Medtronic Percept PC and Oura Ring Gen3, the underlying synchronization strategy is device agnostic. The same general approach could be adapted to other sensing-enabled implanted neurostimulation platforms, including systems such as NeuroPace RNS^20,21^, SceneRay DBS^22,23^, DyNeuMo^24^, and CorTec Brain Interchange^25,26^, so long as implementation accounts for device-specific differences in data access and timestamping. Further, the workflow could be modified to incorporate additional behavioral and physiological measures, including ecological momentary assessments, smartphone-based digital phenotyping, and clinical scores. Integrating these data streams may provide a more comprehensive representation of symptom states during neuromodulation therapy and improve the identification of clinically meaningful biomarkers for next-generation adaptive DBS paradigms.

## CONCLUSION

This work establishes an open-source framework for continuous acquisition and synchronization of wearable-derived behavioral data alongside chronic DBS recordings. We demonstrate reliable temporal alignment across independent device ecosystems and the feasibility of generating longitudinal, behaviorally annotated neural datasets over months of ambulatory monitoring. These datasets enable investigation of state-dependent and temporally structured neural dynamics in the context of objectively measured behavior. More broadly, the framework provides scalable infrastructure for multimodal biomarker discovery and future neurobehaviorally informed neuromodulation strategies.

## Data Availability

All data supporting this study are available online at https://dabi.loni.usc.edu/projects/M7FD26LM28KI.

## ACKNOWLEDGEMENTS

We thank the study participants and their families for their time and involvement in the research. This research was supported by the National Institutes of Health (NIH) National Institute of Mental Health via contract R01MH139889 (N.R.P.), NIH National Institute of Neurological Disorders and Stroke via contract UH3NS136631 (W.K.G., S.A.S., N.R.P., J.A.H.), the Brain & Behavior Research Foundation (N.R.P), and the McNair Foundation (S.A.S., N.R.P.).

